# Risk of Suicide Ideation in Comorbid Substance Use Disorder and Major Depression

**DOI:** 10.1101/2022.02.28.22271669

**Authors:** Vivian N. Onaemo, Timothy O. Fawehinmi, Carl D’Arcy

## Abstract

**Background:** Suicidal behaviour is commonly associated with major depression (MD) and substance use disorders (SUDs). However, there is a paucity of research on risk for suicide ideation among individuals with comorbid SUDs and MD in the general population.

**Objectives:** This study investigated the associated risk of suicide ideation in comorbid SUDs - cannabis use disorder (CUD), alcohol use disorder (AUD), drug use disorder (DUD) with depression in a nationally representative sample.

**Methods:** Multilevel logistic regression models were used to analyze the 2012 Canadian Community Health Survey (CCHS-MHC) data. This is a cross-sectional survey of nationally representative samples of Canadians (n = 25,113) aged 15 years and older residing in the ten Canadian provinces between January and December 2012. Diagnoses of MD episode, AUD, DUD, CUD, and suicide risk were based on the WHO-CIDI-3.0 derived from DSM-IV diagnostic criteria.

**Results:** Comorbidity was found to be the strongest predictor of suicide ideation. Compared to those with no diagnosis, individuals with a comorbid diagnosis of AUD with MDE, CUD with MDE, or DUD with MDE were 9 to 16 times more likely to have suicide ideation. A diagnosis of MDE was a significant predictor of suicide ideation with about a 7-fold increased risk.

**Conclusion:** Suicide is a preventable public health issue. Our study found a significantly increased risk of suicide ideation among persons who have comorbid SUD with MD. Effective integration of mental health and addictions services could mitigate the risk of suicide and contribute to better outcomes.

## Introduction

Globally, suicide is a key public health issue with an estimated 703 000 deaths by suicide annually and even more suicidal attempts (World Health Organization, 2021). In the general population, a prior suicide attempt is considered the single most important predictor of a completed suicide (World Health Organization, 2021). In 2019, suicide was ranked the fourth leading cause of mortality in young people aged 15–29 years worldwide with more than half (58%) of global suicides occurring before the age 50 years (World Health Organization, 2021). Between 2000 and 2019, the age standardized suicide rate increased by 17% in the Americas (World Health Organization, 2021). In high-income countries such as Canada, a history of mental disorders were present in up to 90% of completed suicides (Cavanagh, Carson, Sharpe, & Lawrie, 2003; World Health Organization, 2021).

Substance Use Disorders (SUDs) which includes cannabis use disorder (CUD), alcohol use disorder (AUD), and drug use disorder (DUD) have been found to increase the risk of suicide (Wong, Zhou, Goebert, & Hishinuma, 2013; Westman, et al., 2015; Hjorthoj, et al., 2015; Lynch, et al., 2020). It is estimated that 25 – 50% of all suicides are associated with SUDs and 22% attributable to AUD, meaning that every fifth suicide could have been prevented if alcohol was not consumed (Cavanagh, Carson, Sharpe, & Lawrie, 2003). SUDs are the second most common cause (22.4%) for carrying out suicide in outpatient setting; the increased occurrence in inpatient setting is about two-fold (Driessen, et al., 1998; Park & Kim, 2016; Darvishi, Farhadi, Haghtalab, & Poorolajal, 2015). About 40% of substance dependent individuals that seek treatment have a history of at least one suicide attempt (Yuodelis-Flores & Ries, 2015). Emerging evidence also suggests a role of cannabis on suicidal behavior (Agrawal & Lynskey, 2014; Borges, et al., 2017b; Borges, Benjet, Orozco, Medina-Mora, & Menendez, 2017; Shalit, Shoval, Shlosberg, Feingold, & Lev-Ran, 2016), and cannabis use has been found to be an independent predictor of suicide, with the frequency of use being associated with increased suicide attempts (Schmidt, Tseng, Phan, Fong, & & Tsuang, 2020).

The two most common mental health disorders in North America are SUDs and MD, which often co-exist, especially among treatment-seeking patients (Grant, et al., 2004; Galbaud du Fort, Newman, Boothroyd, & Bland, 1999; RachBeisel, Scott, & Dixon, 1999; Chou, et al., 2012; Grant, et al., 2015; Grant, et al., 2016). A recent systematic review with meta-analysis reported a 3-fold comorbid association between CUD and major depression (Onaemo, Fawehinmi, & D’Arcy, 2021). Suicidal behaviour is commonly associated with depression and alcohol use disorders (World Health Organization, 2014b) and the combination of alcohol dependence and major depression (MD) is considered the leading risk factor for completed suicides (Gliatto & Rai, 1999). While the lifetime risk of suicide is estimated to be 4% in patients with mood disorders (Bostwick & Pankraz, 2000), and 7% in people with alcohol dependence (Schneider, 2009), it increases considerably with comorbidity (Cavanagh, Carson, Sharpe, & Lawrie, 2003). Among adolescents, MD has been found to predict subsequent weekly cannabis use resulting in increased risk for suicidal ideation (Bolanis, et al., 2020). MD and alcohol dependence were responsible for the first and second largest proportion of the suicide disability-adjusted life-years (DALY) that were attributable to mental health and substance use disorders in 2010, respectively (Ferrari, et al., 2014).

Several studies on comorbid SUD and psychiatric disorders have focused on prevalence and risk factors and associations (Brière, Rohde, Seeley, Klein, & Lewinsohn, 2014; Chou, et al., 2012; Grant, et al., 2016) with very few on the impact of that comorbidity such as the associated risk of suicide. Most of the studies that have assessed the impact of comorbid SUD with psychiatric disorders used patient population samples (Britton & Conner, 2010; Glasner-Edwards, et al., 2008; Saatcioglu, Yapici, & Cakmak, 2008; Gliatto & Rai, 1999). In particular, studies on the general Canadian population have not aseesed the impact of comorbid SUD and psychiatric disorders on suicide ideation, thus, a paucity of information (Fuller-Thomson & Hollister, 2016; Fuller-Thomson, West, & Baiden, 2019; Baiden & Fuller-Thomson, 2016; Goodday, Bondy, Sutradhar, Brown, & Rhodes, 2019).

This study aims to the bridge the gap in the literature using a national representative sample of Canadians to assess the impact of comorbid SUD with major depression on the risk of suicide ideation. Since suicide is a sensitive and complex issue, a rare outcome and illegal in some countries, analytical studies on suicides are complicated and suicides are highly likely to be under-reported and misclassified (World Health Organization, 2014b; Borges, Benjet, Orozco, Medina-Mora, & Menendez, 2017). Key risk factors for suicides are suicide attempts and suicide ideation (World Health Organization, 2014b; Nock, et al., 2008), and they, therefore, represent surrogates for studies on suicides and associated factors (Borges, Orozco, Benjet, & Medina-Mora, 2009).

### Objectives

This study aimed to determine the associated risk of suicide ideation in comorbid SUDs - i.e., cannabis use disorder (CUD), alcohol use disorder (AUD), drug use disorder (DUD) with depression in a nationally representative sample.

## Methods

### Subjects

Subjects were participants of the Canadian Community Health Survey (CCHS), 2012: Mental Health Component (CCHS 2012: MH), N= 25,113, response rate = 86.3%. The CCHS 2012: MH provides a comprehensive look at mental health with respect to who is affected by specific mental health disorders, positive mental health, access to and utilization of formal and informal mental health services and support; as well as individual functionality, regardless of the presence of a mental health problem (Statistics Canada, 2014). The survey included persons aged 15 years or more and resident in one of the ten Canadian provinces. Respondents living in certain remote areas, institutions, and reserves were excluded from the survey. In addition, full-time members of the Canadian Forces were not included in this survey. These excluded populations make an estimated 3% of the target national population (Statistics Canada, 2014).

Ethis approval was not sought because this secondary analysis of the CCHS, MH 2012 dataset was done using the Public Use Microdata Files (PUMF). PUMFs are available through University Libraries across Canada. PUMFs are designed to make data widely available while still maintaining confidentiality through aggregating, capping and completely erasing identifying variables (Statistics Canada, 2017b). No consent was required from partcipants to carry out this secondary analysis.

### Measures

#### Major Depression

The World Mental Health version of the Composite International Diagnostic Interview (WMH-CIDI) is a structured diagnostic interview based on symptoms and symptom severity associated with specific psychiatric disorders. The WMH-CIDI algorithm derived from the Diagnostic and Statistical Manual of Mental Disorders IV (DSM IV) was applied to the symptom data to define specific psychiatric diagnosis, in this case, Unipolar Major Depressive Episode (MDE) - lifetime and 12-month (Statistics Canada, 2014).

#### Substance Use Disorders

The WMH-CIDI algorithms derived from DSM IV were applied to the symptom data to define substance use disorders (SUDs), that is, substance abuse and/or dependence for alcohol (alcohol use disorder -AUD), cannabis (Cannabis use disorder – CUD) and other drugs excluding cannabis (drugs use disorder-DUD) in the data. Lifetime and 12-month SUDs were defined (Statistics Canada, 2014).

#### Suicide Ideation

The risk of suicide was assessed using the proxy, suicidal thoughts (ideation). Statistics Canada used the WHM CIDI symptom data to develop algorithms for lifetime and 12-months suicidal thoughts (ideation) in the data. These algorithms classified respondents based on whether they ever (lifetime) thought of committing suicide or taking their own life and whether those thoughts occurred in the past 12 months (Statistics Canada, 2014).

#### Other Measures

Other variables that were included in these analyses were: sociodemographic factors - age, sex, marital status (married, common-law, widowed, divorced or separated, single), highest level of education (less than secondary education, secondary education graduate, some post secondary education, post secondary education graduate), total household income in Canadian dollars (2012) (less than $20,000, $20,000-39,999, $40,000-59,999, $60,000-79,999, $80,000 or more), race (white, non-white), smoking status, personal and family history of mental health disorder, history of a chronic disease and childhood traumatic events (Statistics Canada, 2014).

### Statistical Analysis

A new variable, comorbid SUDs with MDE was first created by merging each SUD (alcohol, cannabis, other drugs) variable and MDE (figure 1). For each SUD, the new variable created had four levels – no diagnosis (neither SUD nor MDE); single diagnosis of SUD (AUD or CUD or DUD); single diagnosis of MDE; and comorbid diagnosis of SUD with MDE. Then, participants were described by sociodemographic characteristics and DSM IV diagnosis.

**Figure 1.**
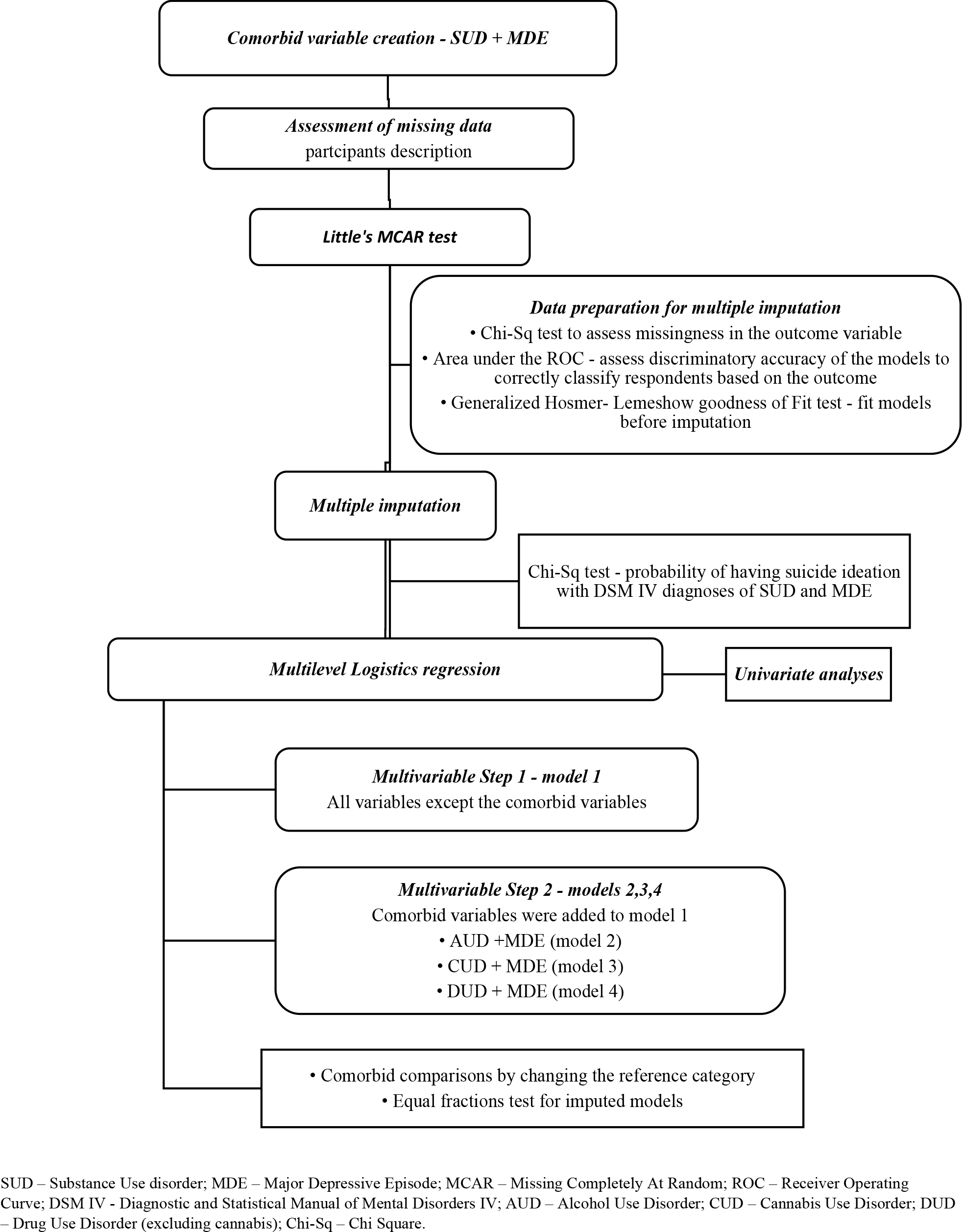
Statistical Analysis Flow Chart.

The Little’s MCAR test on Stata version 14 was used to determine if data was missing completely at random (MCAR) and a justification for a complete case analysis. Data was not missing completely at random (Little’s MCAR test: Prob > chi-square = 0.0000. Multiple imputations was then performed using chained equations following significant Chi Square associations between the independent variables and the missingness in the outcome variable, risk of suicide ideation. Prior to imputation, the area under the receiver operating curve (ROC) was used to assess the discriminatory accuracy of the models to correctly classify respondents based on the outcome. Generalized Hosmer–Lemeshow goodness of fit was used to fit the imputation models prior to imputation. After estimation with the imputed data, the models were tested for equal fraction-missing-information to check that the between-imputation and within-imputation variances were proportional.

Multilevel logistic regression models with province of residence as the group variable were used to assess the outcome –Suicide Ideation. Four models were constructed for the outcome using stepwise multivariate analyses with backward elimination of covariates. In step one, the first model was constructed for the outcome using the sociodemographic factors (age, sex, marital status, education, income, race, smoking), personal and family history of mental illness, history of childhood maltreatment and history of chronic disease. In step two, three other models were constructed by adding the comorbid substance use disorder with major depression variable (alcohol & depression-model 2, cannabis & depression – model 3 and other drugs except cannabis & depression – model 4) to the covariates listed in step one. Since the multilevel models did not allow pairwise comparison post hoc analyses for the variable ‘comorbid substance use disorder with major depression’ in the three comorbid models, comparisons were generated by changing the reference category. The odds ratio (OR) of all covariates in step 1 final model and step 2 final models were reported for the outcome. Confounding was assessed for variables eliminated from the multivariate models and re-entered into the models if present. Since the focus of the analysis was to understand the main effects and the differences between isolated substance use disorders, major depression, and the comorbidities on the risk of suicide ideation, biologically plausible interaction terms such as gender with comorbidity or marital status with comorbidity were not assessed.

A Chi-Square test was done to depict the relationship between the DSM-IV diagnoses (isolated and comorbid) and the probability of having suicide ideation in the population. The complex sampling method of the data was accounted for by using the sampling weights provided by Statistics Canada on the survey (svy) command in Stata. All statistical analyses were carried out using STATA version 14.0.

## Results

### Participants’ characteristics

Table 1 shows weighted percentages of the sociodemographic characteristics and distribution of DSM-IV diagnoses for the target population. A third of the population were residents of Ontario and aged 25-44 years. About half of the population were married, female, were post secondary graduates with total household income of $80,000 or more. Major depressive episode (MDE), Alcohol use disorders (AUD), Cannabis use disorders (CUD), and other drugs excluding cannabis use disorder (DUD) had lifetime prevalence of 11.4%, 19%, 6.8% and 4.0% respectively while the lifetime comorbid diagnoses of MDE with AUD, MDE with CUD, MDE with DUD had prevalence of 3.23%, 1.73% and 1.29% respectively.

**Table 1.**
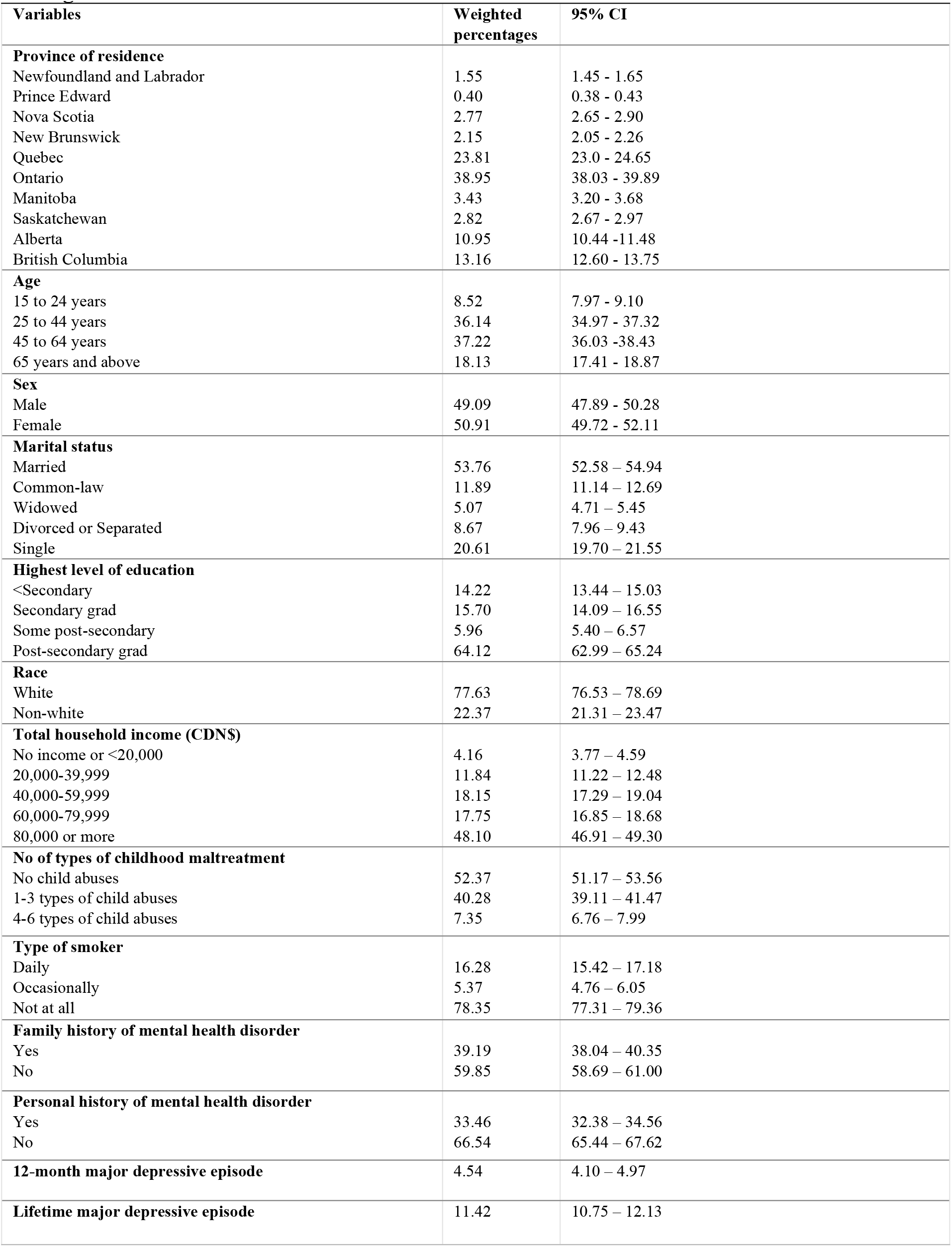
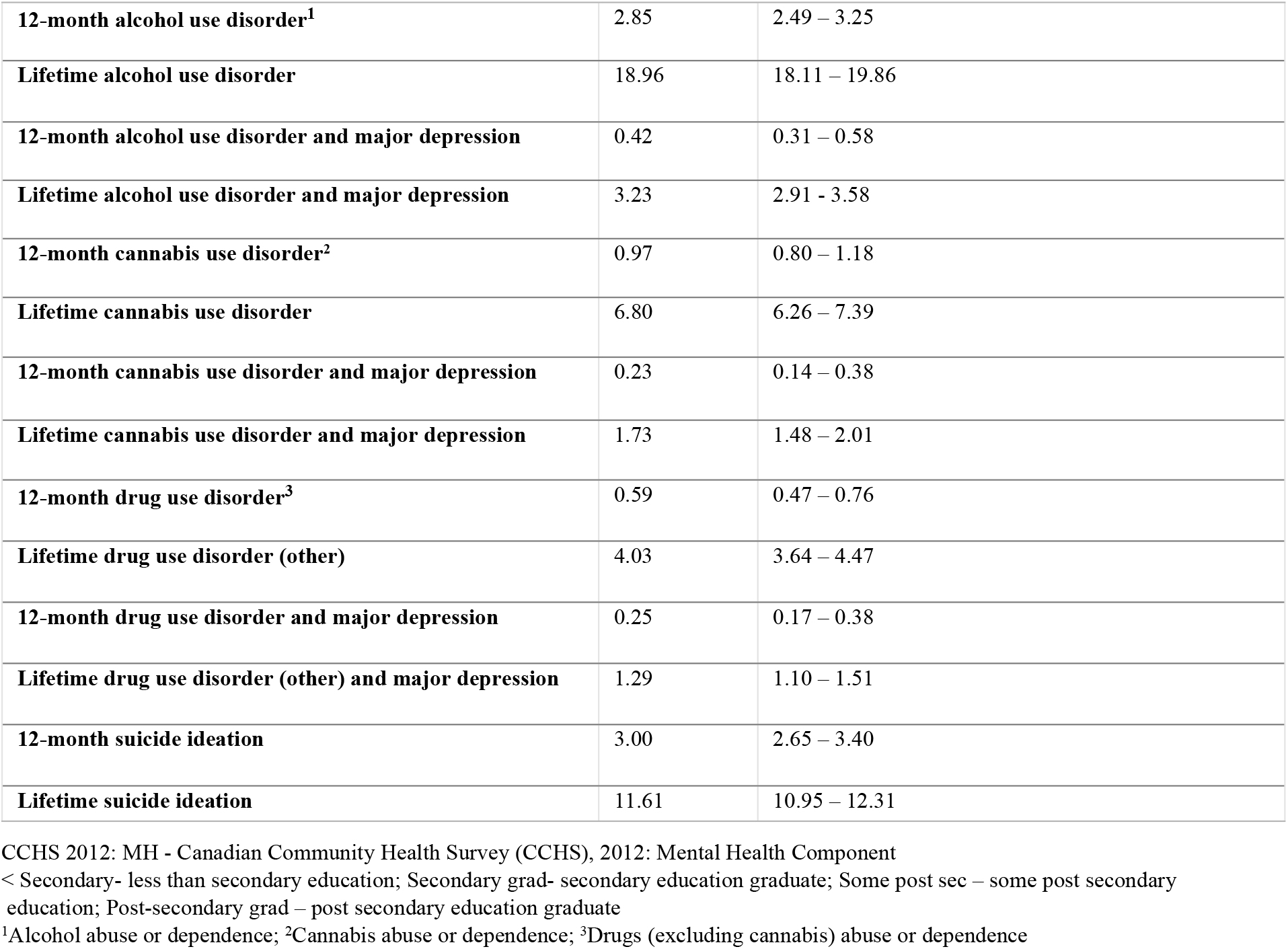
CCHS 2012: MH participants socio-demographic characteristics and distribution of DSM-IV diagnoses.

### Comorbid diagnosis and suicide ideation

The lifetime and 12-month prevalence of suicide ideation were 11.61% and 3.0% respectively (Table 1). Table 2 shows factors associated with suicidal ideation. The odds of having suicidal thoughts is increased among individuals with a history of chronic health condition, childhood maltreatment, family and personal history of mental illness. Older age, being married or common-law, and being a graduate (secondary or post secondary) were protective from suicidal ideation.

**Table 2.** Participants sociodemographic factors associated with 12-month suicide ideation (model 1)

| Crude OR (95% CI) |  | <sup>a</sup>Adjusted |  |
| --- | --- | --- | --- |
|  |  | OR (95% CI) | Statistic (p-value) |
| **Age** |  |  | <0.0001 |
| 15 to 24 years | 1 | 1 |  |
| 25 to 44 years | **0.63** (0.50-0.80)** | **0.61** (0.50-0.75)** |  |
| 45 to 64 years | **0.47** (0.42-0.53)** | **0.39** (0.32-0.49)** |  |
| 65 years and above | **0.22** (0.18-0.27)** | **0.21** (0.16-0.27)** |  |
| **Sex** |  |  |  |
| Male | 1 | 1 |  |
| Female | 1.08 (0.96-1.22) | 1.08 (0.92-1.27) |  |
| **Marital Status** |  |  | 0.0003 |
| Single | 1 | 1 |  |
| Married | **0.37** (0.33-0.41)** | **0.72** (0.64-0.81)** |  |
| Common-law | 0.51 (0.26-1.00) | **0.56** (0.35-0.89)* |  |
| Widowed | **0.36** (0.22-0.60)** | 0.96 (0.56-1.64) |  |
| Divorced or Separated | 0.89 (0.73-1.08) | 1.02 (0.82-1.28) |  |
| **Educational status** |  |  | 0.05 |
| <secondary | 1 | 1 |  |
| Secondary grad | **0.67** (0.52-0.87)** | **0.64** (0.46-0.90)* |  |
| Some post sec | 0.95 (0.65-1.39) | 0.60 (0.34-1.04) |  |
| Post-secondary grad | **0.58** (0.50-0.68)** | **0.69** (0.52-0.91)* |  |
| **Race** |  |  |  |
| White | 1 | 1 |  |
| Non-white | 1.13 (0.88-1.45) | **1.34** (1.12-1.61)** |  |
| **Total Household Income (\$)** | | | <0.0001 |
| No income or <20,000 | 1 | 1 |  |
| 20,000-39,999 | 0.69 (0.44-1.10) | 0.95 (0.64-1.42) |  |
| 40,000-59,999 | **0.48** (0.29-0.80)* | 0.81 (0.52-1.27) |  |
| 60,000-79,999 | **0.44** (0.29-0.65)** | 0.72 (0.46-1.13) |  |
| 80,000 or more | **0.30** (0.22-0.40)** | **0.51** (0.40-0.67)** |  |
| **Type of smoker** |  |  | 0.04 |
| Not at all | 1 | 1 |  |
| Daily | **2.38** (2.09-2.72)** | **1.19** (1.01-1.40)* |  |
| Occasionally | **2.36** (1.48-3.76)** | 1.43 (0.96-2.13) |  |
| **Types of childhood maltreatment** |  |  | <0.0001 |
| None | 1 | 1 |  |
| 1-3 types | **2.66** (1.94-3.64)** | **1.80** (1.21-2.68)* |  |
| 4-6 types | **8.08** (6.08-10.77)** | **3.05** (2.29-4.06)** |  |
| **History of chronic disease** |  |  |  |
| Yes | **3.05** (2.60-3.58)** | **2.47** (2.02-3.03)** |  |
| No | 1 | 1 |  |
| **Family history of mental health disorder** |  |  |  |
| Yes | **2.70** (2.50-2.91)** | **1.62** (1.38-1.89)** |  |
| No | 1 | 1 |  |
| **Personal history of mental illness** |  |  |  |
| Yes | **7.06** (5.61-8.88)** | **4.41** (3.42-5.69)** |  |
| No | 1 | 1 |  |

a – adjusted in a multivariate model; Significant values are marked in bold print
\* p-value <=0.05 \*\* p-value <0.01
Variation explained by the Province of residence is 0.001% (p-value=0.7)
< Secondary- less than secondary education; Secondary grad- secondary education graduate; Some post sec – some post secondary education;
Post-secondary grad – post secondary education graduate

Compared to those with no diagnosis (Table 3), individuals with comorbid diagnosis of AUD with MDE, CUD with MDE, DUD with MDE were 9 to 16 times more likely to have suicide ideation (OR 95% CI - **9.02**, 5.61-14.49**; 11.27**, 7.57-16.78**; 16.19**, 11.10-23.63 respectively). Individuals with comorbid diagnosis of AUD with MDE, CUD with MDE and DUD with MDE had about six to eight-fold increase in the odds of having suicide ideation when compared with individuals with single SUD only (OR 95% CI - **5.58**, 4.28-7.27**; 8.11**, 5.01-13.12**; 6.63**, 5.10-8.64 respectively). A single diagnosis of MDE had about 7-fold increase in the odds of suicide ideation when compared with no diagnosis, and three to five-fold increase when compared with a single diagnosis of DUD, AUD, and CUD (OR 95% CI - **2.92**, 2.18-3.91**; 4.08**, 2.10-7.93**; 5.16**, 3.75-7.09) respectively. A single diagnosis of DUD only, gave a 2-fold increase in the odds of suicide ideation compared to individuals with no diagnosis.

**Table 3.** DSM-IV diagnoses and the risk of 12-month suicide ideation.

| OR (95%CI) |  |  |  |
| --- | --- | --- | --- |
|  | *Ref= ND* | *Ref= SUD* | *Ref= MD* |
| **Alcohol and major depression (model 2)** |  |  |  |
| ND | 1 | 0.62 (0.38-1.02) | **0.15** (0.11-0.22)** |
| AUD only | 1.62 (0.98-2.66) | 1 | **0.25** (0.13-0.48)** |
| MDE only | **6.59** (4.58-9.47)** | **4.08** (2.10-7.93)** | 1 |
| AUD & MDE | **9.02** (5.61-14.49)** | **5.58** (4.28-7.27)** | 1.37 (0.80-2.35) |
| **Cannabis and major depression (model 3)** |  |  |  |
| ND | 1 | 0.72 (0.51-1.01) | **0.14** (0.09-0.22)** |
| CUD only | 1.39 (0.99-1.96) | 1 | **0.19** (0.14-0.27)** |
| MDE only | **7.17** (4.49-11.46)** | **5.16** (3.75-7.09)** | 1 |
| CUD & MDE | **11.27** (7.57-16.78)** | **8.11** (5.01-13.12)** | 1.57 (0.86-2.88) |
| **Other drugs (excluding cannabis) and major depression (model 4)** |  |  |  |
| ND | 1 | **0.41** (0.28-0.60)** | **0.14** (0.09-0.21)** |
| DUD only | **2.44** (1.66-3.59)** | 1 | **0.34** (0.26-0.46)** |
| MDE only | **7.13** (4.76-10.68)** | **2.92** (2.18-3.91)** | 1 |
| DUD & MDE | **16.19** (11.10-23.63)** | **6.63** (5.10-8.64)** | **2.28** (1.47-3.53)** |

Significant values are marked in bold print
\* p-value <=0.05
\*\* p-value <0.01
1 – reference category
Models were adjusted for age, gender, marital status, highest level of education, race, smoking status, types of childhood maltreatment, and family history of mental health disorder in multivariate analyses
Variation explained by the Province of residence: model 1=0.01% (p-value=0.6); model 2=0.01% (p-value=0.4); model 3=0.01% (p-value=0.4)
ND – no diagnosis; AUD - Alcohol use disorders is defined DSM-IV Alcohol Abuse /or Dependence diagnoses.; CUD - Cannabis use disorders is defined DSM-IV Cannabis Abuse /or Dependence diagnoses.; DUD - Drug use disorders is defined as DSM-IV diagnoses of drug abuse/or dependence diagnoses on opiates, sedatives, tranquilizers, amphetamines hallucinogens, heroin, cocaine, inhalants, and/or other drug except cannabis.; MDE - Major Depression is defined DSM-IV diagnosis of major depressive episode. SUD – AUD (model 1), CUD (model 2) and DUD (model 3)

The relationship between DSM-IV diagnosis and the probability of suicide ideation in the population is shown in figure 2. The probability of suicide ideation is <10% in individuals with no diagnosis of either SUD (AUD, CUD or DUD) or MDE. There is 18-34% probability of suicide ideation with isolated diagnosis of a SUD. With a diagnosis of comorbid SUD with MDE, the probability of the suicide ideation ranges from 58-78%.

**Figure 2.**
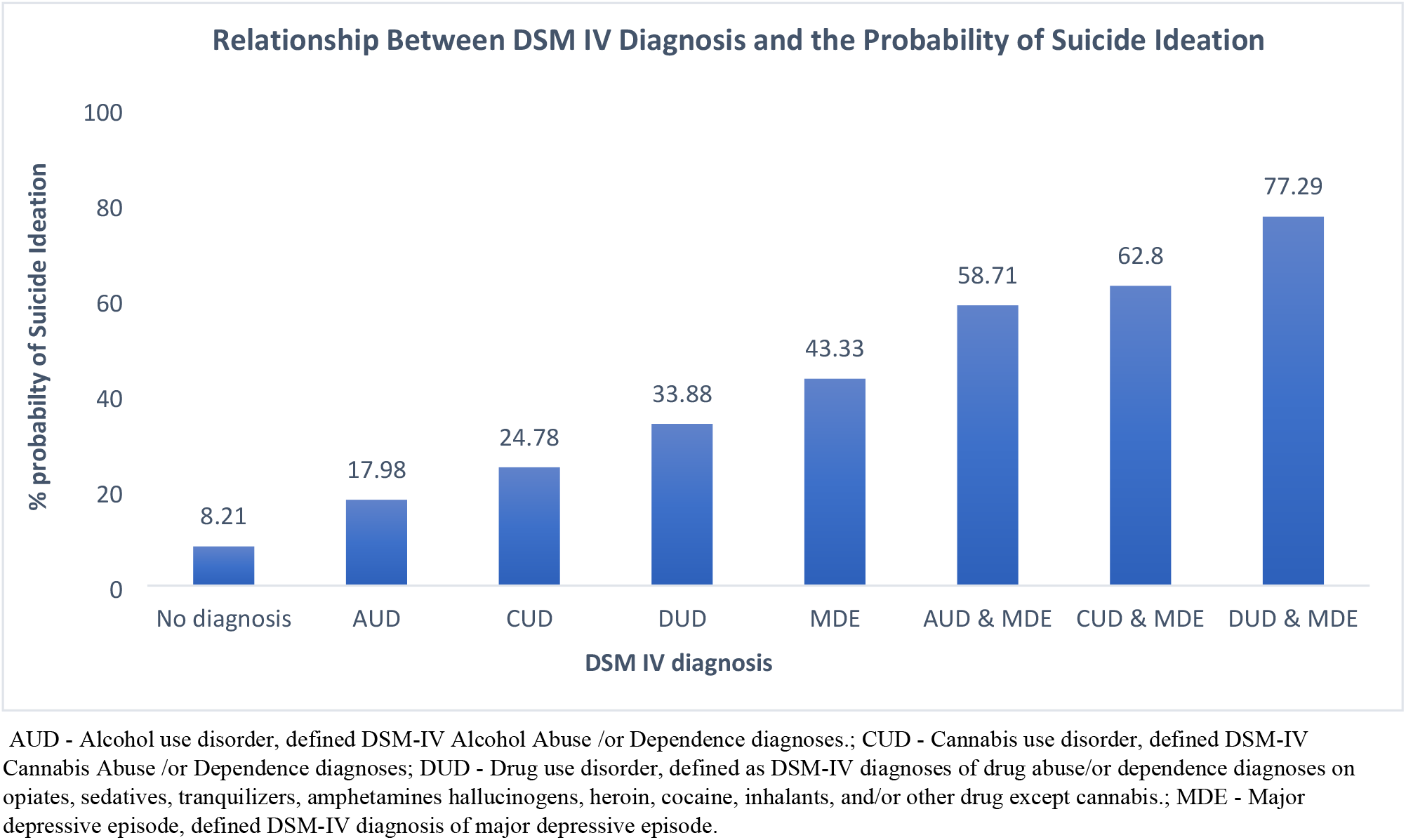
Relationship between DSM-IV diagnosis and the probability of suicide ideation and disability.

## Discussion

### Prevalence and associations of comorbid diagnosis

The past 12-month prevalence of AUD, CUD, DUD, and MDE were 2.9%, 0.97%, 0.6% and 4.5% respectively. The prevalence of MDE, which is about twice that of AUD, is consistent with the findings from a previous survey of the Canadian population (Currie, et al., 2005). In the United States (US) a much higher prevalence was found for depression (Hasin, Goodwin, Stinson, & Grant, 2005), alcohol use disorder (Grant, et al., 2015; Grant, et al., 2004), cannabis and drugs use disorder (Grant, et al., 2016; Grant, et al., 2004) compared to our study. Prevalence of comorbid diagnosis of SUD with MDE in our study was much lower than those found in the US (Grant, et al., 2004). These variations in the prevalence of SUD, MDE and comorbid SUD with MDE could be due to different population characteristics, research methodologies, and diagnostic criteria.

### Comorbid SUD with MDE and risk of suicide ideation

Findings from our study showed a gradual increase from ‘no diagnosis’ to comorbid diagnosis of SUD with MDE in the probability of having suicidal thoughts in the general non-institutionalized population. Our study also showed an increased risk of suicidal thoughts in individuals with comorbid SUD with MDE and sole diagnosis of MDE or DUD, consistent with previous studies (Currie, et al., 2005; Sher, et al., 2008; Cornelius, et al., 1995). This also corroborates other studies that demonstrated AUDs and major depression as the most diagnosed major pathological disorders among persons who commit suicide (Fawcett, Clark, & Busch, 1993; Rudd, Dahm, & Rajab, 1993; Henriksson, et al., 1993).

Our study found that the strongest and most consistent risk factor for suicide ideation was comorbidity. When compared with individuals with no diagnosis, the odds of having suicide ideation was associated with comorbid SUDs with MDE (OR, 95%CI: AUD/MDE - **9.02**, 5.61-14.49; CUD/MDE-**11.27**, 7.57-16.78; DUD/MDE -**16.19**, 11.10-23.63). This risk was much higher than the risk associated with the single diagnosis of MDE (OR, 95%CI - **7.13**, 4.76-10.68). Comorbid SUDs with MDE significantly increased the odds of suicide ideation when compared with the sole diagnosis of AUD (OR, 95%CI -**5.58**, 4.28-7.27), CUD (OR, 95%CI - **8.11**, 5.01-13.12), DUD (OR, 95%CI-**6.63**, 5.10-8.64) and MDE (OR, 95%CI - **2.28**, 1.47-3.53). These findings were consistent with previous findings showing a disproportionate increase in suicide ideation with comorbid AUD with MDE (Britton, et al., 2015; Brière, Rohde, Seeley, Klein, & Lewinsohn, 2014; Cornelius, et al., 1995), and supports findings of increased risk of suicide attempts in SUD co-existing with psychiatric disorders particularly depression (Cottler, Campbell, Krishna, Cunningham-Williams, & Abdallah, 2005; Bakken & Vaglum, 2007; Britton & Conner, 2010). In addition, when compared with the single diagnosis of SUD, a single diagnosis of MDE had a two to five-fold increase in the odds of suicide ideation, also supports previous findings where primary MD without secondary AUD conferred a prospective risk of suicide attempts on the population (Britton, et al., 2015).

This associated increased risk of suicide ideation in comorbid SUDs with MDE could be explained by depression mediating the effect of SUD on suicide or SUD being the consequence of MDE. Studies suggest that suicidal behaviour, aggression, and alcoholism have been linked to abnormally low serotonergic function (Mann & Malone, 1997; Mann, Brent, & Arango, 2001); and this has been proposed to mediate an individual’s genetic and developmental predispositions (Mann, Waternaux, Haas, & Malone, 1999). In addition, studies on humans and animals alike have shown that increased uninhibited psychopathology, substance abuse, and impulsive aggression were secondary to serotonin abnormalities (Crabbe, et al., 1996; Saudou, et al., 1994; Mann, Brent, & Arango, 2001). Substance use disorder have been shown to be associated with the recurrence and persistence of major depression (Onaemo, Fawehinmi, & D’Arcy, 2020) while protracted and untreated depression has been linked to considerable negative outcomes such as increased risk for chronicity or relapse, higher risk for comorbidity and impairments in functioning (Lewinsohn, Rohde, & Seeley, 1998). It is possible that our findings of increased risk of suicide ideation with comorbid SUD with MDE, and isolated MDE may be accounted for by a delay in treatment or non-response to depression treatment.

The significant association of DUD with suicide ideation observed in our study corroborates the earlier studies (Schneider, 2009; Borges, Walters, & Kessler, 2000; Miller, et al., 2011; Petronis, Samuel, Mosckicki, & Anthony, 1990; Borges, Benjet, Orozco, Medina-Mora, & Menendez, 2017). However, in contrast to other previous reports (Sher, et al., 2008; Flensborg-Madsen, et al., 2009; Borges, Benjet, Orozco, Medina-Mora, & Menendez, 2017; Shalit, Shoval, Shlosberg, Feingold, & Lev-Ran, 2016) our study did not find an increased risk of suicide ideation in AUD and CUD only, after controlling for sociodemographic factors and family history of mental illness. This could be attributed to the DSM IV substance use disorder criteria, which entails the presence of DSM IV substance abuse or DSM IV substance dependence without an indication of severity as seen in DSM V. On the other hand, this finding substantiates previous research of no association in low to moderate severity of cannabis use (Shalit, Shoval, Shlosberg, Feingold, & Lev-Ran, 2016).

### Strengths and limitations

A strength of this analysis is that it was based on a nationally representative sample of the Canadian population which provides insight into these comorbidities among individuals irrespective of their health-seeking behaviors. Another strong point was that the diagnoses of SUD and MDE were derived from DSM IV criteria using a structured diagnostic interview and algorithm (WMH-CIDI). This study included different comparison groups of SUD and MDE highlighting the actual effects of comorbid associations. Missing values were accounted for using multiple imputations and the complex data structure was accounted for with multilevel random effects models and survey weights, enabling generalizability. Limitations of this study include the cross-sectional study design which does not allow for causal inference. Further studies would be required to assess causality. Individuals living on reserves and other Aboriginal settlements, in institutions and full-time members of the Canadian Forces which make up about 3% of the total population were excluded from the data. Therefore, our analysis may have underestimated the true strength of associations of interest.

## Conclusion and Public Health Implications

This study does provide further evidence of associated risk of suicide ideation among persons who have comorbid alcohol use disorders, cannabis use disorders, drug use disorders with major depression. Suicide ideation among individuals with MD and comorbid SUDs tend to present with more severe mood symptoms, higher risk of suicide attempts, poor functioning, more psychiatric comorbidities, and increased mortality (Blanco, et al., 2012; Gadermann, Alonso, Vilagut, Zaslavsky, & Kessler, 2012). Despite this knowledge, public health mental services are seriously underutilized by individuals with suicidal thoughts (Luoma, Martin, & Pearson, 2002), and their suicide ideations usually do not come to the attention of clinicians (Bruffaerts, et al., 2011). In addition, evidence suggests increased suicides among clients with AUD or SUDs who have associated high disengagement from services in the form of skipping scheduled appointments and leaving the hospital after declining further treatment (Hunt, et al., 2006).

Effective integration of mental health and addictions services with a collaborative approach that includes a multi-disciplinary team of MHA treatment providers, healthcare practitioners, family members, and community resources are essential to the effective engagement, development of preventative strategies and potentially overcoming a number of structural and behavioural impediments that support suicide ideation. This may contribute to better treatment outcomes.

## Data Availability

Access to the data is available to bona fide researchers through institutions participating in Statistics Canada Data Liberation Initiative (DLI) including university libraries throughout Canada? see https://www.statcan.gc.ca/eng/dli/dli. Access can be arrange directly through DLI enquiries.

## Author Contributions

Conceived and designed the study: VNO

Analyzed the data: VNO

Contributed materials/analysis tools: VNO, TOF, CD.

Wrote the paper: VNO, TOF, CD.

## Acknowledgements

The data that support the findings of this study are from the Public Use Microdata File (PUMF) of the Canadian Community Health Survey – Mental Health Component, Statistics Canada survey #5015.

Access to the data is available to bona fide researchers through institutions participating in Statistics Canada Data Liberation Initiative (DLI) including university libraries throughout Canada – see https://www.statcan.gc.ca/eng/dli/dli.

Access can be arrange directly through DLI enquiries.

